# Selection of appropriate reference genes for normalization of qRT-PCR based gene expression analysis in SARS-CoV-2, and Covid-associated Mucormycosis infection

**DOI:** 10.1101/2022.03.15.22272441

**Authors:** Sunil Kumar, Ayaan Ahmad, Namrata Kushwaha, Sheetal Negi, Kamini Gautam, Anup Singh, Pawan Tiwari, Rakesh Garg, Richa Agarwal, Anant Mohan, Anjan Trikha, Alok Thakar, Vikram Saini

## Abstract

Selection of reference genes in quantitative PCR is critical to determine accurate and reliable mRNA expression. Despite this knowledge, not a single study has investigated the expression stability of housekeeping genes to determine their suitability to act as reference gene in SARS-CoV-2 or Covid19 associated mucormycosis (CAM) infections. Herein, we address these gaps by investigating the expression stability of nine most commonly used housekeeping genes including *TBP, CypA, B2M, 18S, PGC-1α, GUSB, HPRT-1, β-ACTIN* and *GAPDH* in the patients of varying severity (asymptomatic, mild, moderate and severe). We observed significant differences in the expression of candidate genes across the disease spectrum. Next, using various statistical algorithms (delta Ct, Normfinder, Bestkeeper, RefFinder and GeNorm), we observed that *CypA* demonstrated the most consistent expression across the patients of varying severity and emerged as the most suitable gene in Covid19, and CAM infections. Incidentally, the most commonly used reference gene *GAPDH* showed maximum variations and found to be the least suitable. Lastly, comparative evaluation of expression of *NRF2* is performed following normalization with *GAPDH* and *CypA* to highlight the relevance of an appropriate reference gene. Our results reinforce the idea of a selection of housekeeping genes only after validation especially during infections.

## Introduction

Understanding of the changes in expression profile of selected genes is critical to develop diagnostics, therapeutic, and prognostics approaches against infectious agents (1-6). In this context, real-time quantitative polymerase chain reaction (qRT-PCR) is often the preferred method to monitor relative changes in gene expression due to high sensitivity, specificity and broad quantification range for a high throughput and accurate expression profiling. A major challenge, towards appropriate harnessing of powerful tool of qRT-PCR, however, is the necessity of having an endogenous control or reference gene from the same sample to ensure an accurate assessment of target gene’s expression. The prerequisite for a suitable reference gene not only warrants an adequate and consistent expression in the sample of interest, but more importantly, the reference gene should show minimal variability in the expression between samples, and under experimental conditions used for investigation. Nonetheless, it is widely known that the preferred reference genes may often demonstrate significant variability in expression compromising the outcomes, and data quality rendering even inaccurate results (7-9). Therefore, appropriate validation of reference genes in any new experimental system is of utmost importance.

Housekeeping genes due to their constitutive and stable expression often qualify as reference genes in qRT-PCR studies (10-12). Indeed, Glyceraldehyde-3-phosphate dehydrogenase (*GAPDH*), hypoxanthine guanine phosphoribosyl transferase (*HPRT-1*), cyclophilin A (*CypA*), ribosomal protein L13A (*RPL13A*) and *β-ACTIN* are among one of the most commonly used housekeeping genes used as reference gene/ internal controls for qRT-PCR analysis (13-14). For viral infections including Covid19, *β-ACTIN, GAPDH*, and *18S* are popularly used as internal controls for normalization (4,15-18). However, most of these genes do not consistently manifest stable expression under various experimental conditions (19-21). This could be especially critical in the context of SARS-CoV-2 infection since it manifests a wide spectrum of disease severity ranging from asymptomatic, mild, moderate and severe; each with a distinct clinical features/parameter (22-24). Moreover, during SARS-CoV-2 infection or following recovery, there can be acute co-infections especially that of fungal (mucormycosis) which may further impact the stability and expression of the appropriate reference gene. Notably, over 45,000 people in India have developed Covid-associated mucormycosis (CAM) signifying it as one of the major Covid19 associated co-infection. CAM has a fatality rate of >50% and was declared an epidemic in India by health care authorities (25). Unfortunately, despite the high mortality of CAM and need to develop better diagnostics and therapeutic tools, there is not a single study focusing on identifying the appropriate reference gene and investigating the impact of CAM on commonly used reference genes. Likewise, to the best of our knowledge, there is hardly any study which has looked into the expression stability of housekeeping genes in SARS-CoV-2 or Covid19 infection to determine their suitability to act as internal reference gene.

Therefore, it is clear that an unsuitable reference gene may compromise or even cause mis-identification of key genes involved in disease severity, target identification and associated outcomes during infection. Moreover, the acute onset of deadly fungal infections of mucormycosis along with or post-recovery from SARS-CoV-2, further necessitates the evaluation of impact of these co-infections on the stability and expression of the appropriate internal control. In this study, we have addressed these knowledge gaps by evaluating the gene expression stability of nine widely used housekeeping genes in PBMCs isolated from patients having spectrum of Covid19 severity, and also patients that developed CAM. Our results clearly show that significant variations exist in the expression of housekeeping genes during SARS CoV2 or CAM infection, and selects the most appropriate candidate reference gene establishing the utility of the study.

## Material and Method

### Patients’ selection and Cohort

Blood samples from different age and sex group ranging from 25-60 years (at least 8 patients for each group) of SARS-CoV-2 infected (mild, moderate, severe, and asymptomatic) and Covid-associated mucormycosis i.e., pre-surgery (Pre-CAM) and post-surgery (Post-CAM) along with healthy control (Total subjects=56, **Table 1**). We collected blood samples at the time of admission of patients to the clinic by taking informed consent. Individuals in healthy category were Covid19 negative as confirmed by qRT-PCR, with no past history of Covid19 and comorbidities (diabetes, flu, cough, TB, etc.). This study was approved by institutional Human Ethics committee All India Institute of Medical Sciences (AIIMS), New Delhi (IEC-435/02.07.2021, RP-34/2021 and IEC-419/02.07.2021).

**Table 1.** Description of Subjects enrolled in study cohort. Patient cohort of 56 individuals (39 males and 17 females) enrolled from different groups (healthy, asymptomatic, mild, moderate, severe, pre-CAM, and post-CAM) along with their comorbidity status.

|  |  |  |
| --- | --- | --- |
| **Individual enrolled** | 56<br>*Median age (42)* |  |
| **Gender distribution** | *Male*<br>39 | *Female*<br>17 |
| **Age (range in years)** | 17-75 | 16-70 |
| **Median Age (years)** | 45 | 35.5 |
| **Diabetes** | 16 | 3 |
| **Hypertension** | 2 | 1 |
| **Anaemia** | 1 | 0 |
| **Hyperthyroidism** | 1 | 1 |
| **Malignancy** | 1 | 0 |
| **Heart disease** | 1 | 0 |

### Criteria of disease and clinical severity

All the cases of Covid19 and CAM were clinically and microbiologically confirmed at AIIMS, New Delhi. Severity of Covid19 infection was ascertained as per the institutional clinical definitions as shown below:

a. Mildly symptomatic: Patients with proven SARS-CoV-2 infection having mild symptoms, i.e., fever and cough and/or shortness of breath with respiratory rate > 24/min and SpO_2_ > 94%
b. Moderately Sick: Patients with proven SARS-CoV-2 infection having moderate disease, i.e., fever with cough and /or shortness of breath with respiratory rate > 24/min and SpO_2_< 94%.
c. Severe patients: Patients with proven SARS-CoV-2 infection having moderate to severe disease, i.e., respiratory failure requiring mechanical ventilation and clinical evaluation by the attending physician.
d. Asymptomatic: Patients with proven SARS-CoV-2 infection but without any apparent symptoms.
e. Covid-associated Mucormycosis (CAM): Microbiologically confirmed mucormycosis in patients having Covid19 or immediate after recovery from Covid19 ascertained by RT-PCR report.
f. Post-surgery patients Covid-associated Mucormycosis (Post-CAM): These patients were having CAM, and reported in the hospital at advance stage of infection or were referred from other medical care centers. As per the clinical requirement, these patients have to undergo surgery immediately and were still under treatment for CAM at the time of sample collection.

The design of study did not influence the clinical care or course of treatment for any of the patient.

### Isolation of Peripheral blood mononuclear cells (PBMCs) from blood

PBMCs were isolated from the patient blood as described earlier (26). Briefly, blood samples were collected from each subject and PBMCs were isolated using density gradient centrifugation (Ficoll™ HiMedia, India). Buffy coat containing PBMCs was removed carefully, and washed twice with PBS at 1500rpm for 10 min. PBMCs were counted under microscope and approximately 10^8^ cells were transferred to 1mL TRIzol (Invitrogen, USA) containing microfuge tubes for RNA isolation followed by downstream processing including qRT-PCR.

### RNA isolation and cDNA synthesis

The RNA was extracted from PBMCs using TRIzol reagent as per well-established protocols (1,3,23). RNA concentration (ng/mL) and purity (A260/A280) were determined spectroscopically (Cyatation Biotek, USA). cDNA synthesis was carried out using the iScript™ cDNA synthesis kit (Bio-Rad, USA). Briefly, 1µg RNA, 4µL of 5x iScript™ reaction mix, 1µL of iScript™ reverse transcriptase and nuclease-free water was added together to a final volume of 20µL and incubated at 5 min at 25°C for priming followed by reverse transcription for 20 min at 46°C. Finally, to inactivate the enzyme, samples were incubated at 95°C for 1 min followed by a hold on 4°C. Once cDNA is synthesised, it was diluted 10 times and the resulting RNA was used for real-time PCR amplification.

### Selection of candidate genes, primer design and optimization of qRT-PCR conditions

From literature review, we selected nine most frequently used housekeeping genes namely *TBP, CypA, B2M, 18S, PGC-1α, GUSB, HPRT-1, β-ACTIN*, and *GAPDH* to determine their suitability to act as a reference gene in Covid-19 infection and CAM (**Table 2**). In addition to these housekeeping genes, we also used nuclear factor erythroid 2-related factor 2 (*NRF2*) to perform evaluation of changes in gene expression pattern of this gene based on the reference gene identified from our study. We used molecular biology database (NCBI) and online software such as Primer 3 and NCBI-BLAST (https://blast.ncbi.nlm.nih.gov/Blast.cgi) for extraction of gene sequences and primer designing. For our study, we designed primers that were 18 to 24 base pairs in length; a Tm of 50°C to 60°C and GC content ranging from 40-60%. NCBI-BLAST (https://blast.ncbi.nlm.nih.gov/Blast.cgi) was carried out to confirm primer specificity prior to synthesizing. Primer specificity was validated using qRT-PCR based melting curve analysis. We used SYBR Green-based assay to quantify gene amplification in CFX Opus 96 qRT-PCR machine (BioRad, USA). All the reactions were performed at least in duplicates with the final volume for each reaction was 20µL with the following components: 10µL of iTaq™ Universal SYBR Green supermix (2X), 2µL of diluted cDNA template, 0.5µL of forward and reverse primer (10µM) and nuclease-free water to the final volume. The reaction was programmed for an initial denaturation at 95°C for 5 min, followed by 39 cycles of denaturation at 95°C for 10s, annealing at 60°C for 10s, and extension at 72°C for 30s. The melting curve was obtained by heating the amplicon from 65-95°C in a continuous acquisition mode.

**Table 2.**
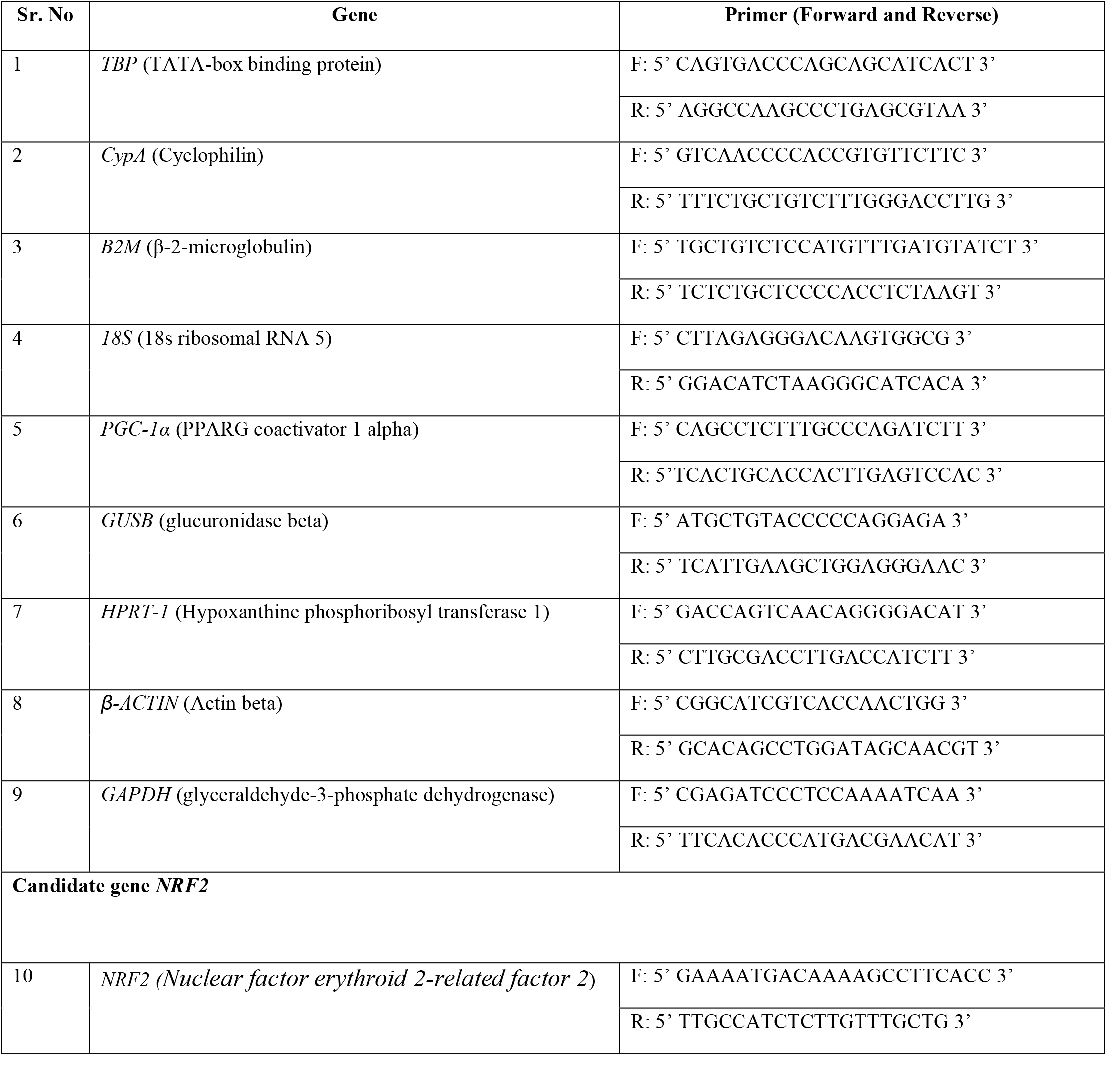
Primer sequences of candidate reference genes used in this study.

### Statistical Analysis and the application of algorithms for the selection of the most suitable reference gene

Statistical analysis was performed using modules available in GraphPad ver 5.01. For software analysis, statistical tool in-built into the programs were used in the default mode. The significance of differences in *NRF2*-fold change was ascertained by using a Mann Whitney paired t-test (*, *P-value* <0.05; **, *P-value* < 0.001; ***, *P-value* < 0.0001 and ‘ns’ represents non-significant). Data represents mean ± standard error of the mean (SEM) performed at least twice in duplicates.

Different statistical tools namely delta Ct, Bestkeeper, Normfinder, GeNorm, and RefFinder to calculate stability of each reference gene across the disease’s severity. All these software/programs have inbuilt statistical tools to score for variations and decide on the most preferred genes. After predicting stability of individual reference genes with each tool, a final comprehensive gene stability analysis was also determined for each gene among different severity levels using RefFinder algorithm.

## Results

### Melt curve analyses indicate specificity of the gene primers

Using clinically confirmed cases of Covid19 and CAM, we isolated PBMCs and obtained a high purity RNA as ascertained by 260/280 ratio (Supplementary Table 1). We selected total of ten genes for this study, which includes the nine most frequently used housekeeping genes namely *TBP, CypA, B2M, 18S, PGC-1α, GUSB, HPRT-1, β-ACTIN, and GAPDH* (4,17) (Table 2). *NRF2* was chosen as a candidate test gene to evaluate differences in expression profile *vis a vis* reference gene of interest. Following the synthesis of cDNA, we first determined whether PCR amplicons for the individual genes are of expected size. We observed a single amplification product for each primer pair as evident by presence of a single peak in the melting curve analysis (**Figure 1**). This highlighted that there is no non-specific binding of primer pairs and therefore primers are suitable for further downstream applications.

**Figure 1:**
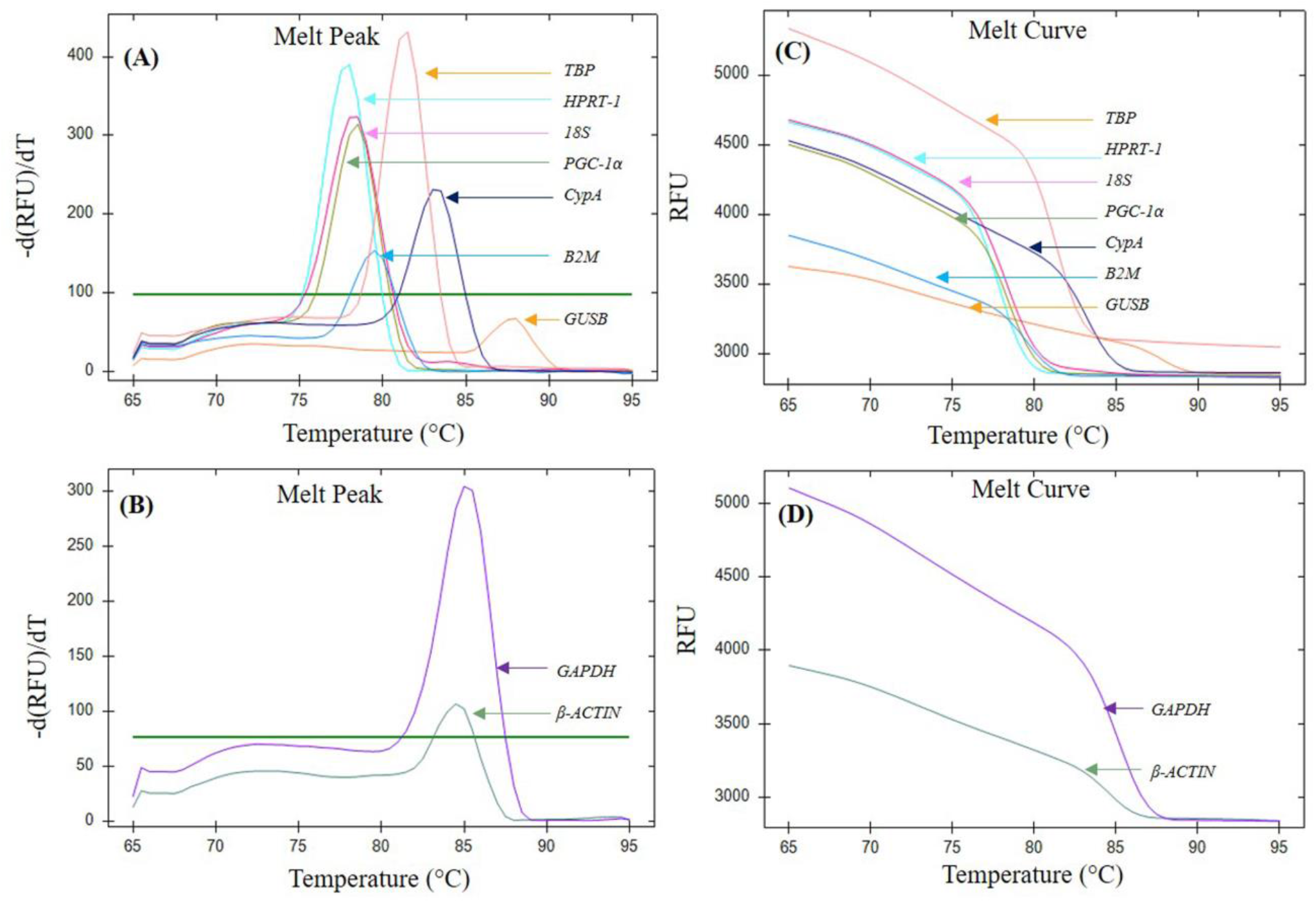
Assessment of dissociation characteristics of candidate reference genes using melt curve analysis. Melt peak of (A) *TBP, CypA, B2M, 18S, PGC-1α, NRF-2, GUSB* and *HPRT-1*; and (B) *GAPDH* and *β-ACTIN*. Melt curve of (C) *TBP, CypA, B2M, 18S, PGC-1α, GUSB* and *HPRT-1*; and (D) *GAPDH* and *β-ACTIN*. A single Melt peak and a single melt curve for each individual gene indicates the generation of a single amplicon following primer amplification, respectively (RFU=Relative Fluorescence Unit).

### Analysis of candidate reference genes reveals heterogeneity in gene expression in Covid19 and CAM patients

We next investigated the changes in the expression of candidate housekeeping genes in patients (*n=56*) belonging to the following clinically defined groups (total=7); healthy, asymptomatic, mild, moderate, severe, pre-CAM (CAM patients prior to surgery) and post-CAM (CAM patients’ post-surgery). First of all, the selected nine candidate reference genes were within the dispersal range of Ct value (range >6.43-39.69); and also fold increase in PCR product per cycle i.e., amplification efficiency was within the accepted range i.e. (90-100%).

Based on the disease severity of SARS CoV-2 infection or associated fungal infection, we noticed that significant differences exist in the expression of candidate reference genes within the group as well as among the groups across the disease spectrum (**Figure 2**). Surprisingly, the most commonly used internal control gene including in Covid19 studies i.e., *GAPDH*, showed significant variation within healthy subjects, and also between healthy and Covid19 subjects with a moderate or severe or CAM infections. For *β-ACTIN*, we noticed that expression of this gene was comparable in healthy, mild and asymptomatic individuals but substantial variations could also be observed in individuals with moderate or severe or CAM infection. Likewise, analysis of *PGC-1α*, another commonly used housekeeping gene also demonstrated significant variations among patients having asymptomatic or CAM infection.

**Figure 2:**
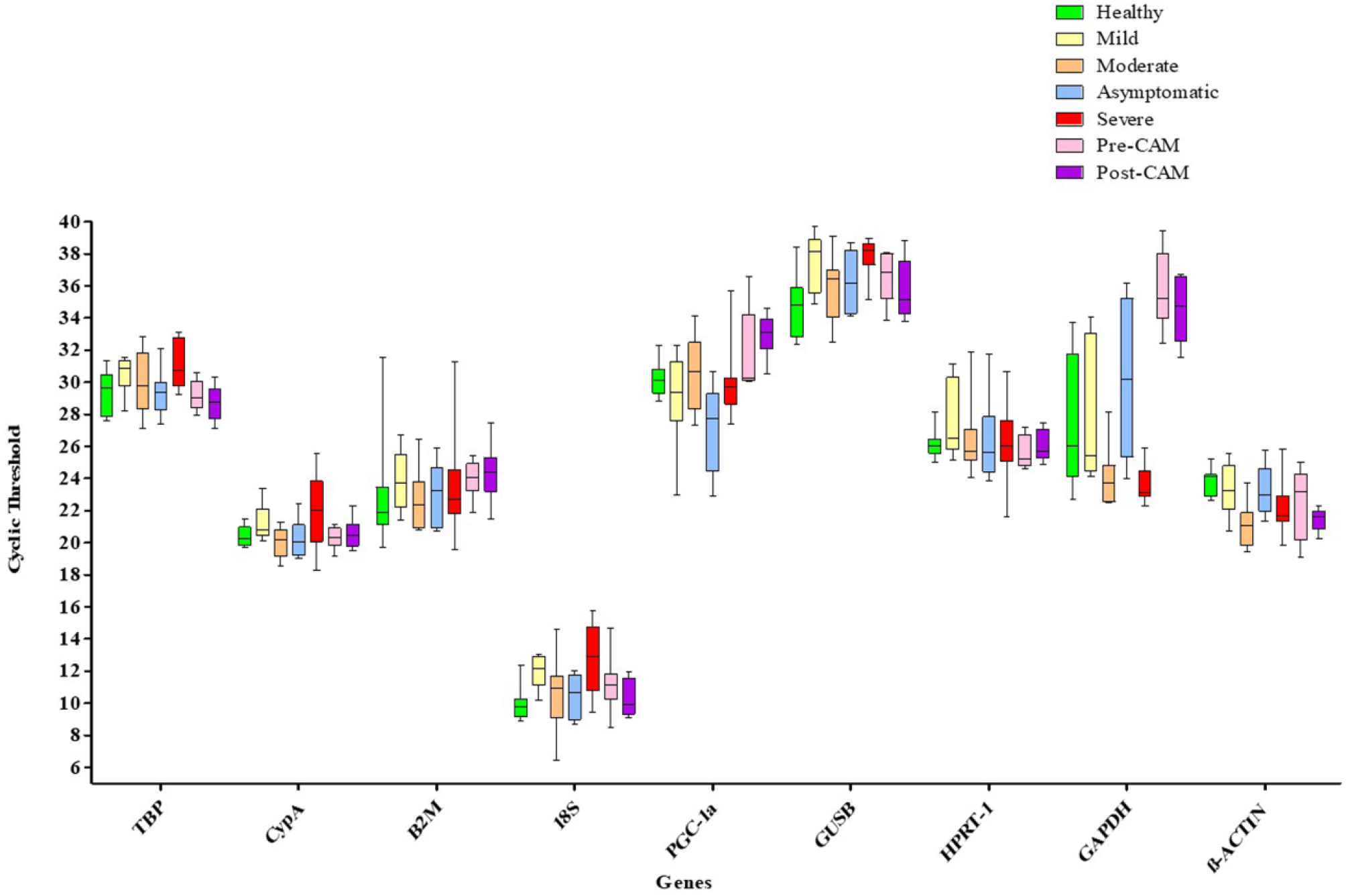
Analysis of candidate reference genes reveals significant heterogeneity across Covid19 patients of varying severity. Boxplot of mean Ct values (Y-axis) for candidate reference genes (X-axis) used in this study is indicated. The line across the box represents median Ct value, whereas whiskers indicate range of Ct values obtained in a particular group of patients. Data represents a total of 56 subjects (n=56, male-39; female-17; 8 subjects/group) including the healthy individuals.

For *18S* and *GUSB*, even though the range of Ct values were comparable across the groups, these genes consistently expressed either very early (*18S*, 6.43-15.73; 95% CI range -10.56-11.53) or very late (*GUSB*, 32.40 - 39.69; 95% CI range - 35.88-36.94) across all groups as indicated by their respective Ct values. Ideally, for an internal reference gene, a moderate expression level as exemplified by Ct value (15.0-30.0) is essential in order to accurately represent the quantitative expressions of the genes of interest (27). As shown in **figure 2**, only *TBP, CypA, B2M* and *HPRT-1* have a comparable mean Ct value across SARS CoV-2 disease spectrum and CAM infection. Nonetheless, which one of these genes would be the most suitable to use as an internal reference gene during Covid19 infection and/or CAM is not immediately obvious. For an objective analysis, we would require algorithms employing statistical methods to select the most suitable reference gene across various groups.

### Application of statistical algorithms identifies *CypA* and *TBP* as the most appropriate reference genes for SARS-CoV-2 infection and associated mucormycosis

Considering the significant variation in Ct values observed among patients inter-or intra-group, it was prudent to apply statistical tools to determine the appropriate and reliable reference gene. In this context, we have employed popular algorithms such as comparative delta-Ct method, Bestkeeper, GeNorm, RefFinder and Normfinder for determining the stability of reference genes. These algorithms are programmed to capture various aspects of expression data including gene stability, intra- and/or inter-group variations, standard deviations etc. to indicate a suitable reference gene.

Using comparative delta-Ct method, we observed that *GAPDH* and *PGC-1α* showed the highest standard deviations across different groups of Covid19 and CAM infections as is also evident by the cumulative standard deviation data for individual genes (**Figure 3A**). *CypA* has the least average and the least cumulative standard deviation in expression followed by *18S* and *TBP*. Another tool Bestkeeper on the other hand, indicated *PGC-1α* and *β-ACTIN* as the two most stable reference genes. We also used popular tools namely Normfinder and GeNorm to investigate the suitability of candidate genes as internal reference genes. Both of these tools revealed *GAPDH* to be the least suitable gene based on the highest arithmetic mean of all pairwise variations (indicated by M value in GeNorm), and the highest stability value which is inversely related to stability of an internal reference gene (i.e., the most stable gene have the lowest stability value). Both of these tools identified *CypA* as the most suitable internal reference gene followed by *18S*. Remarkably, none of the two most popularly used internal control genes in Covid19 i.e., *GAPDH* and *β-ACTIN* showed stability of expression across the patient pool of varying severity. Further, none of the above tools used in this study consensually identified ideal reference gene even though majority algorithms predicted *CypA* as the most suitable candidate reference gene across the disease spectrum (**Figure 3**).

**Figure 3:**
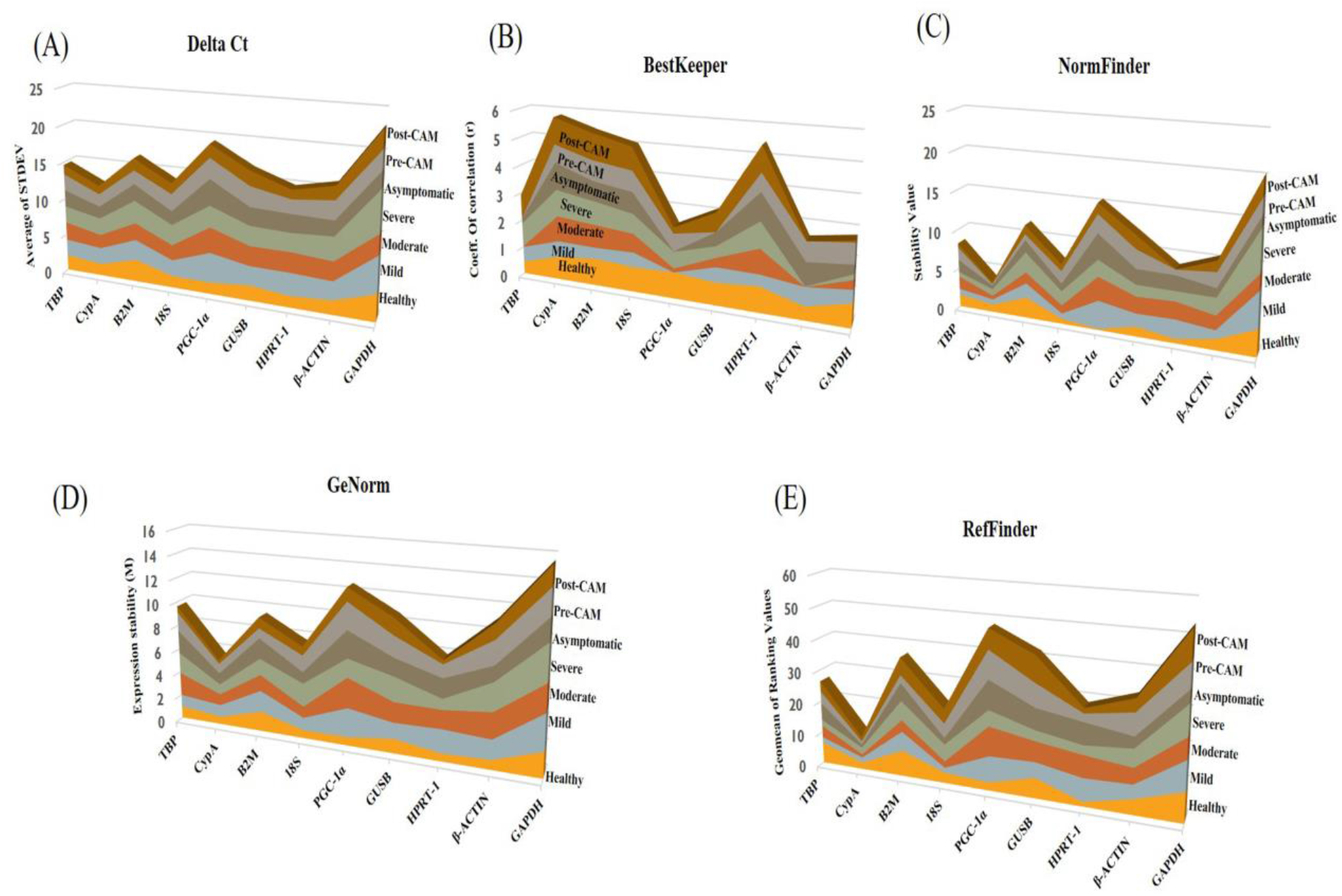
Comparison of candidate reference genes based on cumulative ranking score using different statistical algorithms. (A) Intra-group average standard deviations for individual patient groups were obtained using delta Ct method, and are added to obtain a cumulative average standard deviation (Y-axis). The highest cumulative average standard deviation (i.e., *GAPDH*) indicates the least gene stability (X-axis). (B) The graph represents a cumulative correlation coefficient obtained by adding intra-group correlation coefficients (r) (Y-axis) using Bestkeeper algorithm. The candidate gene yielding the lowest cumulative value (i.e., *PGC-1α*) indicates the highest stability. (C) In Normfinder algorithm, intra-group stability value (Y-axis) based on coefficient of variations are added to obtain a cumulative stability value. The candidate gene yielding lowest cumulative stability value (i.e., *CypA*) indicates the most stable reference gene. (D) In GeNorm algorithm, intra-group expression stability (M value) based on the average pairwise variation of an individual gene with all other genes used as control to obtain a cumulative expression stability (Y-axis). The gene yielding the highest M value (i.e., *GAPDH*) indicates the lowest stability. (E) In RefFinder algorithm, intra-group geometric mean (Y-axis) based on geometric mean of ranking values calculated by all above algorithms are added to obtain a cumulative geometric mean of ranking values. The gene yielding the lowest cumulative geometric mean of ranking values (i.e., *CypA*) indicates the highest stability.

We next applied a widely used algorithm RefFinder (28) that considers the output of other algorithms (comparative delta-Ct method, Bestkeeper, GeNorm, and Normfinder) as input data to yield a final comprehensive ranking based on geometric mean (GM) of ranking values. Based on RefFinder analysis, *CypA* emerged as the topmost suitable gene as it yielded the least cumulative GM (10.35) [GM has an inverse relation with gene stability, lower the GM-higher the stability]. This value is more than two times less than the second and third most suitable candidate reference gene identified in this fashion i.e., *18S* (GM=22.92) and *TBP* (GM=26.43), respectively. On the other hand, *GAPDH* emerged as the least suitable reference gene as evident by the highest cumulative GM (53.51) which is over five times higher than *CypA. β-ACTIN*, the other popular and widely used reference gene in Covid19 and other viral infections, could only be ranked 5^th^ for its suitability to act as reference gene out of the nine candidate reference genes evaluated in this study (Supplementary Table 2).

### Evaluation of *NRF2* gene expression in Covid19 and CAM infections highlights the relevance of selection of an appropriate reference gene

After identifying *CypA* as the most suitable reference gene for Covid19, and CAM infections, we next evaluated the relevance of having an appropriate internal control by using nuclear factor erythroid 2-related factor 2 (*NRF2)* gene expression as an example. *NRF2* is a master regulator for an array of antioxidant responses and regulates pathophysiological conditions including/during infections. Since role of oxidative stress as an important hallmark of Covid19 pathophysiology is well documented (29), we expected *NRF2* expression to be a good candidate for this study. Using RNA sequencing, it was recently shown that *NRF2* is suppressed in Covid19 patients and that activating *NRF2* expression could be a viable therapeutic strategy (30).

We evaluated whether *NRF2* expression in Covid19 and CAM patients would vary based on the choice of the internal reference gene used for qRT-PCR analysis. For normalization of expression, we used *CypA*, the most suitable reference gene based on our results, and *GAPDH*, the commonly used reference gene in Covid19. As is evident from **figure 4**, *GAPDH* could not accurately capture the apparently large heterogeneity of the Covid19 spectrum, and CAM patients. Contrary to the existing knowledge, we observed a significantly higher induction of *NRF2* in asymptomatic, mild, moderate, Pre-CAM and Post-CAM groups when *GAPDH* was used as internal control instead of *CypA* (**Figure 4**). Only in severely sick, there was no apparent difference in gene expression of *NRF2* based on normalization with *CypA* or *GAPDH*. Clearly, the normalization of *NRF2* with *GAPDH* would rather be misleading and won’t able to accurately represent broad clinical spectrum of Covid19 and CAM and possibility of *NRF2* activation-based therapies proposed by several independent studies (17,18,30). Our study therefore highlights the relevance of an appropriate selection of internal control for the normalization of qRT-PCR data.

**Figure 4:**
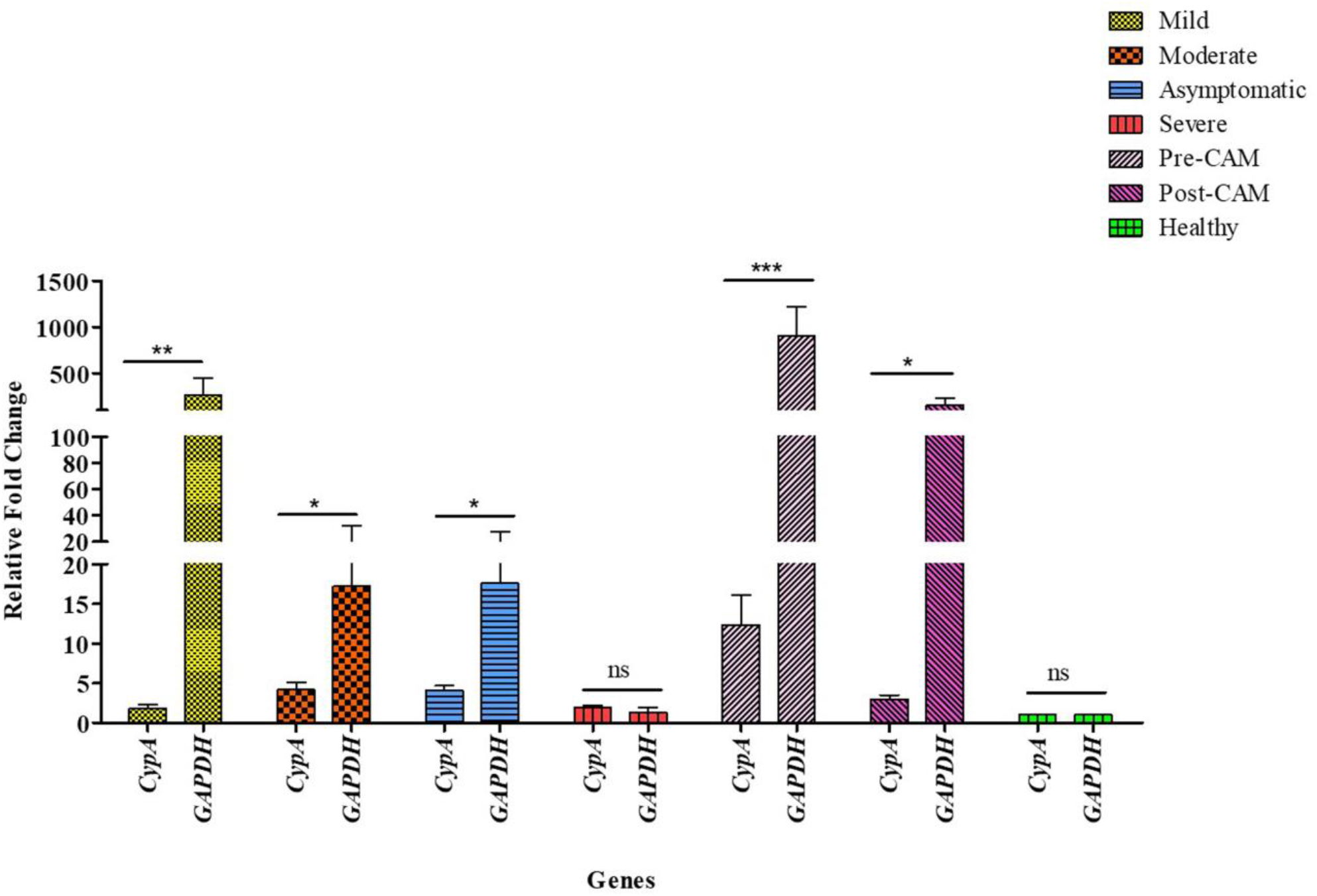
Comparative analysis of expression profile of *NRF2* based on the usage of *GAPDH* or *CypA* as internal control gene. The analysis of differences in fold change in the *NRF2* expression (Y-axis) reveals significant differences based on data normalization *vis à vis GAPDH* and *CypA*, the least and the most stable reference genes from our study, respectively. Healthy subjects, represents the baseline *NRF2* expression. Data represents mean ± standard error of the mean (SEM) performed with at least eight subjects in duplicates. Significance is ascertained by using a Man-Whitney paired t-test (*, *P-value* <0.05; **, *P-value* < 0.001; ***, *P-value* < 0.0001, and ns=non-significant).

## Discussion

The availability of qRT-PCR instrumentation in standard molecular biology laboratories and medical diagnostics centers even in the developing and low-income countries affords unique opportunities to study gene expression profiles that can recognize, differentiate, and classify discrete gene subsets to predict disease outcome, or response to therapy. Study of key gene families, and pathways across different geographies and genotypes will help in the development of improved therapeutics for an effective disease management. As indicated earlier, reference genes are widely used as internal controls to accurately interpret expression data. Unfortunately, there is no universal internal reference gene and an evaluation of internal reference genes to ascertain their suitability is advisable at the onset of a new condition.

Both Covid19 infection and CAM are relatively less explored with respect to their molecular mechanisms to develop better management and treatment strategies. Covid19 has so far impacted 404,910,528 peoples with 5,783,776 deaths in more than 191 countries including low middle income countries (LMIC) and developing countries (WHO coronavirus dashboard dated 11^th^ Feb, 2022). Likewise, for CAM incidence rate varies from 0.005 to 1.7 per million population, whereas, India alone has 140 per million population as per WHO. Unfortunately, though despite the need to understand human responses upon onset of Covid19 and CAM at the transcriptional level, a formal standardization of reference genes for the qRT-PCR analysis of gene expression in Covid19 and mucormycosis infected patients has not been reported. Especially, it is not clear how spectrum of the disease severity differentially impacts the expression of housekeeping genes.

In this study, we evaluated nine housekeeping genes commonly used for normalization of gene expression profiles to determine ideal reference gene during infection of Covid19, and CAM. These genes were selected as they are expressed extensively in all cells and have different functions, in order to avoid over representation of genes belonging to same biological pathway. Firstly, PBMCs are used in this study as they are one of the most accessible and easy to work with cell types. Further, gene expression profile in PBMCs correlates well with the systemic gene expression changes in the diseases condition (31). Both these features make them an attractive candidate to investigate differential gene expressions, particularly for quantitative traits involved in the phenotypic outcomes (32).

*GAPDH* and *β-ACTIN* are one of the most widely used reference genes for performing gene expression analysis by qRT-PCR in different human cells including in Covid19 studies. Importantly, in case of CAM, there is not a single gene expression study that has been performed so far. Based on our analysis, GAPDH emerged as the least suitable reference gene as apparently significant variation in its expression exists in patients of varying severity. Another gene used in the Covid19 study *β-ACTIN* also was found unsuitable as it could only be ranked 5^th^ out of the nine-candidate analyzed in this study for their ability to act as reference gene. So, our results reinforce the idea of proper validation of internal reference genes before using them to avoid making erroneous observations. The other candidate genes like *18S* had very low, and *GUSB* had very high expression levels as compared with other housekeeping genes in both healthy and diseased condition making them unsuitable as reference genes for qRT-PCR analysis. Based on our results, *CypA* has the most stable expression across varying spectrum of Covid19 infection including in the patients having infection of mucormycosis. Consequently, *CypA* emerged as the most appropriate candidate to be used as a reliable reference gene in qRT-PCR-based analysis of Covid19 and mucormycosis for normalization of gene expression. Although, *18S* emerged as the second most stable gene but it is expressed very early, while a medium-high level expression is desirable for an ideal reference gene (27). This may limit its utility as a potential reference gene in qRT-PCR based relative gene expression analysis. On the other hand, *TBP* consistently showed a moderate expression across the Covid19 spectrum and has only a marginal difference between its cumulative geometric means (26.43) *vis a vis 18S* (22.92) making it a better choice than *18S*.

It is well known that selection of unsuitable reference genes may weaken the detection sensitivity of the target genes, and even result in inaccurate results (33). We also observed that the normalization of *NRF2* gene expression yielded significantly different results when expression was normalized with *CypA* or *GAPDH*, the most suitable and the least suitable genes respectively, identified in this study. Normalization with *GAPDH* highlighted significant expression heterogeneity (2-fold to over 900-fold) across different spectrum of Covid19 and CAM infection. Out of conditions tested in this study, significant up-regulation of *NRF2* was observed in mild (>250 fold), asymptomatic (>15 fold), moderate (>12 fold), pre-CAM (>900 fold), and post-CAM (>150 fold). This is in contrast to the RNA sequencing studies, which have shown that *NRF2* expression is extremely suppressed in Covid19 and independent groups have argued for therapeutic strategies that could stimulate *NRF2* expression (30,34). Evidently, *NRF2* expression appears to fluctuate drastically when data is normalized with *GAPDH* as reference gene and rather creates the impression of over-expression of *NRF2*, a master regulator gene, as a major problem.

On the other hand, use of *CypA* as the internal control in the same patients yielded a stable picture of *NRF2* induction in mild (<2 fold), asymptomatic (<5 fold), moderate (<7 fold), pre-CAM (<13 fold), and post-CAM (<3 fold). Only in case of severe group, we obtained similar data from normalization with *GAPDH* and *CypA* (<2-fold induction in both cases). Therefore, it is clear that only normalization with *CypA*, the most suitable reference gene identified in our study, could accurately capture the heterogeneity of infection and yielded very stable expression across the conditions.

To the best of our knowledge, the current investigation is the only systematic analysis of stability of housekeeping genes during the course of infection of Covid19, and mucormycosis. The results clearly establish that an expression of housekeeping gene may vary due to infection and reinforces the idea of careful selection of housekeeping genes post-validation.

## Data Availability

All data produced in the present study are available upon reasonable request to the authors.

## Acknowledgment

We would like to thank staff of Laboratory of Infection Biology and Translational Research namely Krishan Pal, Shailendra Kumar, Nidhi, and Shalini Sharma for providing assistance in laboratory work, and with sample collection and processing. We are also thankful to all the subjects and study volunteers for their participation to this study. We thank Bio-Rad India for providing RT-PCR instrumentation available to us during the pandemic.

## AUTHOR CONTRIBUTIONS

*Concept, design and overall supervision: VS; Clinical sample processing: SK, NK, SN, KG; Data Acquisition: SK and AA, Analysis and interpretation of data: SK, AA and VS; Sample Collection and Clinical inputs: AS, PT, RG, RA, AM, AT, and ALT; Drafting of the manuscript: SK and VS wrote and edited the manuscript, all authors provided inputs/comments on the manuscript; Obtained funding: VS*.

## Data availability

All data are available from the corresponding authors upon request.

## References

1. Saini, V., Cumming, B.M., Guidry, L., Lamprecht, D.A., Adamson, J.H., Reddy, V.P., Chinta, K.C., Mazorodze, J.H., Glasgow, J.N., Richard-Greenblatt, M., et al. (2016) Ergothioneine Maintains Redox and Bioenergetic Homeostasis Essential for Drug Susceptibility and Virulence of Mycobacterium tuberculosis. Cell Reports, 14, 572–585.

2. Reddy, V.P., Chinta, K.C., Saini, V., Glasgow, J.N., Hull, T.D., Traylor, A., Rey-Stolle, F., Soares, M.P., Madansein, R., Rahman, M.A., et al. (2018) Ferritin H Deficiency in Myeloid Compartments Dysregulates Host Energy Metabolism and Increases Susceptibility to Mycobacterium tuberculosis Infection. Front. Immunol., 9, 860.

3. Saini, V., Chinta, K.C., Reddy, V.P., Glasgow, J.N., Stein, A., Lamprecht, D.A., Rahman, Md.A., Mackenzie, J.S., Truebody, B.E., Adamson, J.H., et al. (2020) Hydrogen sulfide stimulates Mycobacterium tuberculosis respiration, growth and pathogenesis. Nat Commun, 11, 557.

4. Manne, B.K., Denorme, F., Middleton, E.A., Portier, I., Rowley, J.W., Stubben, C., Petrey, A.C., Tolley, N.D., Guo, L., Cody, M., et al. (2020) Platelet gene expression and function in patients with COVID-19. Blood, 136, 1317–1329.

5. Kumar, P., Saini, K., Saini, V. and Mitchell, T. (2021) Oxalate Alters Cellular Bioenergetics, Redox Homeostasis, Antibacterial Response, and Immune Response in Macrophages. Front. Immunol., 12, 694865.

6. McClain, M.T., Constantine, F.J., Nicholson, B.P., Nichols, M., Burke, T.W., Henao, R., Jones, D.C., Hudson, L.L., Jaggers, L.B., Veldman, T., et al. (2021) A blood-based host gene expression assay for early detection of respiratory viral infection: an index-cluster prospective cohort study. The Lancet Infectious Diseases, 21, 396–404.

7. Glare, E. M., Divjak, M., Bailey, M. J., & Walters, E. H. (2002). β-Actin and GAPDH housekeeping gene expression in asthmatic airways is variable and not suitable for normalising mRNA levels. Thorax, 57(9), 765–770.

8. Liu, L.-L., Zhao, H., Ma, T.-F., Ge, F., Chen, C.-S. and Zhang, Y.-P. (2015) Identification of Valid Reference Genes for the Normalization of RT-qPCR Expression Studies in Human Breast Cancer Cell Lines Treated with and without Transient Transfection. PLoS ONE, 10, e0117058.

9. Zhao, Y., Wong, L. and Goh, W.W.B. (2020) How to do quantile normalization correctly for gene expression data analyses. Sci Rep, 10, 15534.

10. Turabelidze, A., Guo, S. and DiPietro, L.A. (2010) Importance of housekeeping gene selection for accurate reverse transcription-quantitative polymerase chain reaction in a wound healing model: Accurate RT-qPCR in a wound healing model. Wound Repair and Regeneration, 18, 460–466.

11. Chapman, J.R. and Waldenström, J. (2015) With Reference to Reference Genes: A Systematic Review of Endogenous Controls in Gene Expression Studies. PLoS ONE, 10, e0141853.

12. Piazza, V., Bartke, A., Miquet, J. and Sotelo, A. (2017) Analysis of Different Approaches for the Selection of Reference Genes in RT-qPCR Experiments: A Case Study in Skeletal Muscle of Growing Mice. IJMS, 18, 1060.

13. Cherin, T., Catbagan, M., Treiman, S. and Mink, R. (2006) The effect of normothermic and hypothermic hypoxia–ischemia on brain hypoxanthine phosphoribosyl transferase activity. Neurological Research, 28, 831–836.

14. Ahn, K., Huh, J.-W., Park, S.-J., Kim, D.-S., Ha, H.-S., Kim, Y.-J., Lee, J.-R., Chang, K.-T. and Kim, H.-S. (2008) Selection of internal reference genes for SYBR green qRT-PCR studies of rhesus monkey (Macaca mulatta) tissues. BMC Mol Biol, 9, 78.

15. Kuchipudi, S.V., Tellabati, M., Nelli, R.K., White, G.A., Perez, B.B., Sebastian, S., Slomka, M.J., Brookes, S.M., Brown, I.H., Dunham, S.P., et al. (2012) 18S rRNAis a reliable normalisation gene for real time PCR based on influenza virus infected cells. Virol J, 9, 230.

16. Geyer, H., Bauer, M., Neumann, J., Lüdde, A., Rennert, P., Friedrich, N., Claus, C., Perelygina, L. and Mankertz, A. (2016) Gene expression profiling of rubella virus infected primary endothelial cells of fetal and adult origin. Virol J, 13, 21.

17. Ouyang, Y., Yin, J., Wang, W., Shi, H., Shi, Y., Xu, B., Qiao, L., Feng, Y., Pang, L., Wei, F., et al. (2020) Downregulated Gene Expression Spectrum and Immune Responses Changed During the Disease Progression in Patients With COVID-19. Clinical Infectious Diseases, 71, 2052–2060.

18. Atwan, Z.W. (2020) GAPDH spike RNA as an alternative for housekeeping genes in relative gene expression assay using real-time PCR. Bull Natl Res Cent, 44, 32.

19. Chu, K., Jung, K.-H., Kim, S.-J., Lee, S.-T., Kim, J., Park, H.-K., Song, E.-C., Kim, S.U., Kim, M., Lee, S.K., et al. (2008) Transplantation of human neural stem cells protect against ischemia in a preventive mode via hypoxia-inducible factor-1α stabilization in the host brain. Brain Research, 1207, 182–192.

20. Arien-Zakay, H., Lecht, S., Bercu, M.M., Tabakman, R., Kohen, R., Galski, H., Nagler, A. and Lazarovici, P. (2009) Neuroprotection by cord blood neural progenitors involves antioxidants, neurotrophic and angiogenic factors. Experimental Neurology, 216, 83–94.

21. Sarnowska, A., Braun, H., Sauerzweig, S., Reymann, K.G. and Minhas, G. The neuroprotective effect of bone marrow stem cells is not dependent on direct cell contact with hypoxic injured tissue.

22. Gavriatopoulou, M., Korompoki, E., Fotiou, D., Ntanasis-Stathopoulos, I., Psaltopoulou, T., Kastritis, E., Terpos, E. and Dimopoulos, M.A. (2020) Organ-specific manifestations of COVID-19 infection. Clin Exp Med, 20, 493–506.

23. Saini, V., Kalra, P., Sharma, M., Rai, C., Saini, V., Gautam, K., Bhattacharya, S., Mani, S., Saini, K. and Kumar, S. (2021) A Cold Chain-Independent Specimen Collection and Transport Medium Improves Diagnostic Sensitivity and Minimizes Biosafety Challenges of COVID-19 Molecular Diagnosis. Microbiol Spectr, 9, e01108–21.

24. Mehta, O.P., Bhandari, P., Raut, A., Kacimi, S.E.O. and Huy, N.T. (2021) Coronavirus Disease (COVID-19): Comprehensive Review of Clinical Presentation. Front. Public Health, 8, 582932.

25. Mishra, Y., Prashar, M., Sharma, D., Akash Kumar, V.P. and Tilak, T.V.S.V.G.K. (2021) Diabetes, COVID 19 and mucormycosis: Clinical spectrum and outcome in a tertiary care medical center in Western India. Diabetes & Metabolic Syndrome: Clinical Research & Reviews, 15, 102196.

26. Patel, M., Yarlagadda, V., Adedoyin, O., Saini, V., Assimos, D.G., Holmes, R.P. and Mitchell, T. (2018) Oxalate induces mitochondrial dysfunction and disrupts redox homeostasis in a human monocyte derived cell line. Redox Biology, 15, 207–215.

27. Li, J., Zhang, Z., Xu, C., Wang, D., Lv, M. and Xie, H. (2020) Identification and validation of reference genes for real-time RT-PCR in Aphelenchoides besseyi. Mol Biol Rep, 47, 4485–4494.

28. Gorji-Bahri, G., Moradtabrizi, N., Vakhshiteh, F. and Hashemi, A. (2021) Validation of common reference genes stability in exosomal mRNA-isolated from liver and breast cancer cell lines. Cell Biol Int, 45, 1098–1110.

29. Delgado-Roche, L. and Mesta, F. (2020) Oxidative Stress as Key Player in Severe Acute Respiratory Syndrome Coronavirus (SARS-CoV) Infection. Archives of Medical Research, 51, 384–387.

30. Olagnier, D., Farahani, E., Thyrsted, J., Blay-Cadanet, J., Herengt, A., Idorn, M., Hait, A., Hernaez, B., Knudsen, A., Iversen, M.B., et al. (2020) SARS-CoV2-mediated suppression of NRF2-signaling reveals potent antiviral and anti-inflammatory activity of 4-octyl-itaconate and dimethyl fumarate. Nat Commun, 11, 4938.

31. Mesko, B., Poliska, S. and Nagy, L. (2011) Gene expression profiles in peripheral blood for the diagnosis of autoimmune diseases. Trends in Molecular Medicine, 17, 223–233.

32. Lam, L.L., Emberly, E., Fraser, H.B., Neumann, S.M., Chen, E., Miller, G.E. and Kobor, M.S. (2012) Factors underlying variable DNA methylation in a human community cohort. Proceedings of the National Academy of Sciences, 109, 17253–17260.

33. Köhsler, M., Leitsch, D., Müller, N. and Walochnik, J. (2020) Validation of reference genes for the normalization of RT-qPCR gene expression in Acanthamoeba spp. Sci Rep, 10, 10362.

34. McCord, J.M., Hybertson, B.M., Cota-Gomez, A., Geraci, K.P. and Gao, B. (2020) Nrf2 Activator PB125® as a Potential Therapeutic Agent against COVID-19. Antioxidants, 9, 518.

