## Supplementary Table 1 and 2 for "Selection of appropriate reference genes for normalization of qRT-PCR based gene expression analysis in SARS-CoV-2, and Covid-associated Mucormycosis infection"

### Supplementary Tables

| Healthy | | Mild | | Moderate | | Asymptomatic | |
|---|---|---|---|---|---|---|---|
| *Sample Code* | *260/280 ratio* | *Sample Code* | *260/280 ratio* | *Sample Code* | *260/280 ratio* | *Sample Code* | *260/280 ratio* |
| *H1* | 1.938 | *A006* | 2.021 | *B003* | 1.900 | *D010* | 2.010 |
| *H2* | 1.995 | *A008* | 2.125 | *B004* | 2.000 | *D011* | 2.025 |
| *H3* | 1.970 | *A010* | 2.040 | *B011* | 1.924 | *D012* | 2.000 |
| *H4* | 2.001 | *A012* | 1.900 | *B012* | 2.013 | *D013* | 1.980 |
| *H5* | 1.990 | *A013* | 1.800 | *B013* | 1.967 | *D015* | 1.841 |
| *H6* | 2.004 | *A027* | 1.978 | *B010* | 2.020 | *D017* | 1.500 |
| *H7* | 2.100 | *A028* | 2.046 | *B015* | 2.100 | *D024* | 2.041 |
| *H8* | 1.900 | *A044* | 1.991 | *B001* | 1.890 | *D035* | 2.030 |
| **Severe** | | **Pre-CAM** | | **Post-CAM** | | | |
| *Sample Code* | *260/280 ratio* | *Sample Code* | *260/280 ratio* | *Sample Code* | *260/280 ratio* | | |
| *C006* | 1.929 | *CAM03* | 1.700 | *CAM13* | 1.690 | | |
| *C012* | 1.622 | *CAM8* | 1.980 | *CAM15* | 1.670 | | |
| *NC001* | 1.711 | *CAM16* | 1.735 | *CAM34* | 1.850 | | |
| *NC002* | 1.824 | *CAM17* | 1.640 | *CAM37* | 1.810 | | |
| *NC005* | 1.725 | *CAM19* | 1.920 | *CAM38* | 1.710 | | |
| *NC008* | 1.824 | *CAM24* | 1.800 | *CAM4* | 1.900 | | |
| *NC009* | 1.400 | *CAM28* | 1.590 | *CAM6* | 1.826 | | |
| *NC011* | 1.792 | *CAM47* | 1.940 | *CAM10* | 1.849 | | |

**Supplementary Table 1: Description of RNA quality (260/280) of samples.**

| Genes | Healthy | Mild | Moderate | Severe | Asymptomatic | Pre-CAM | Post-CAM | Cumulative Geomean | Comprehensive Ranking |
|---|---|---|---|---|---|---|---|---|---|
| *CypA* | 2.00 | 1.73 | 1.00 | 2.21 | 1.00 | 1.00 | 1.41 | 10.35 | **1** |
| *18S* | 3.22 | 1.57 | 2.00 | 5.23 | 2.00 | 4.47 | 4.43 | 22.92 | **2** |
| *TBP* | 6.48 | 1.86 | 3.72 | 1.97 | 5.42 | 3.76 | 3.22 | 26.43 | **3** |
| *HPRT-1* | 1.41 | 7.00 | 6.73 | 2.82 | 6.65 | 2.45 | 1.57 | 28.63 | **4** |
| *β-ACTIN* | 4.61 | 4.23 | 4.79 | 5.47 | 3.41 | 6.70 | 3.98 | 33.19 | **5** |
| *B2M* | 8.00 | 6.00 | 3.57 | 6.05 | 5.48 | 2.45 | 5.44 | 36.99 | **6** |
| *GUSB* | 6.19 | 4.73 | 6.19 | 3.25 | 6.51 | 7.14 | 8.00 | 42.01 | **7** |
| *PGC-1α* | 2.51 | 8.00 | 9.00 | 4.90 | 9.00 | 8.74 | 6.19 | 48.34 | **8** |
| *GAPDH* | 9.00 | 9.00 | 6.40 | 9.00 | 3.87 | 7.24 | 9.00 | 53.51 | **9** |

**Supplementary Table 2: Conclusive comprehensive ranking based on geometric means (GM) of ranking values using RefFinder analysis.**
